# How can spectrum bias impact expectations of test performance: a secondary modeling study estimating novel swab-based test outcomes across populations

**DOI:** 10.64898/2026.07.02.26357020

**Authors:** Sarah Zaidi, Tushar Garg, Luan Nguyen Quang Vo, Melissa Sander, Andrew James Codlin, Rachel Louise Byrne, Vibol Iem, Sayera Banu, John Bimba, Victor Santana Santos, Cyrille Mbuli, Han Thi Nguyen, Augustine Choko, Steve Wandiga, Stephen John, Bertie Squire, Tom Wingfield, Jacob Creswell

**Affiliations:** Stop TB Partnership, Geneva, Switzerland; Friends for International TB Relief, Hanoi, Viet Nam; Department of Global Public Health, Karolinska Institutet, Stockholm, Sweden; Center for Health Promotion and Research (CHPR), Bamenda, Cameroon; Centre for Tuberculosis Research, Departments of Clinical Sciences and International Public Health, Liverpool School of Tropical Medicine, Liverpool, UK; Department of Clinical Sciences, Liverpool School of Tropical Medicine, Liverpool, UK; icddr,b, Dhaka, Bangladesh; Zankli Research Centre, Bingham University, Karu, Nigeria; Department of Medicine, Federal University of Sergipe, Lagarto, Brazil; FIT RD Social Enterprise Company Limited (FIT RD), Hanoi, Viet Nam; Division of Infectious Diseases, Department of Medicine, Karolinska Institutet, Stockholm, Sweden; Malawi Liverpool Wellcome Research Programme, Blantyre, Malawi; Departments of Clinical Sciences and International Public Health, Liverpool School of Tropical Medicine, Liverpool, UK; Kenya Medical Research Institute (KEMRI), Nairobi, Kenya; Janna Health Foundation, Jimeta Yola, Adamawa State, Nigeria; Tropical and Infectious Diseases Unit, Liverpool University Hospitals NHS Foundation Trust, Liverpool, UK

## Abstract

**Background:** New swab-based near-point-of-care (NPOC) tests offer a potentially lower-cost, simpler alternative to Xpert MTB/RIF Ultra (Xpert-Ultra) testing for tuberculosis (TB) diagnosis. However, it is critical to understand how their performance may differ across diverse populations to inform programmatic rollout and real-world clinical decision-making.

**Methods:** We modeled the performance of testing sputum swabs with the MiniDock MTB assay and the Xpert MTB/RIF (Xpert) using positive percentage agreement (PPA) with Xpert-Ultra, disaggregated by semi-quantitative grade. PPA for MiniDock MTB was derived from a random-effects meta-analysis of three diagnostic accuracy studies. PPA for Xpert was derived from data from an early diagnostic study. PPA estimates were applied to 1,248 positive Xpert-Ultra test results from Phase 1 of the Start4All study across seven countries, disaggregated by facility-based (n=1,033) and community-based (n=215) participant recruitment, comparing a single overall PPA to an Xpert Ultra semi-quantitative grade-stratified model (from Trace to High).

**Results:** Pooled overall PPA was 84.8% (95% CI: 67.1–93.8%) for MiniDock MTB and 93.1% (90.3–95.2%) for Xpert. Grade-stratified modeling revealed lower PPAs at ‘Very Low’ and ‘Trace’ semi-quantitative grades: MiniDock MTB 56.5% and 33.8%; Xpert 66.7% and 23.1%, respectively. When grade-stratified estimates were applied to the Start4All data, both tests performed similarly, missing 241/1,248 and 228/1,248 positive Xpert-Ultra results, respectively. The single-value model overestimated performance most markedly in community settings, predicting 10.8% and 19.1% more positive results than the grade-stratified model for MiniDock MTB and Xpert, respectively.

**Conclusion:** MiniDock MTB sputum swabs perform comparably to Xpert when assessed using grade-stratified modeling. Both tests are likely to miss a greater proportion of people with Xpert-Ultra positive results in community settings, where paucibacillary disease is more common. Using single point estimates for diagnostic accuracy can substantially overestimate real-world performance, highlighting the importance of evaluating diagnostics across the full spectrum of TB disease.

## Introduction

Tuberculosis (TB) remains a pressing public health problem, killing more people than any other infectious disease. High mortality is driven by 2.4 million people who develop TB annually who are not detected, treated, and reported.(1) The World Health Organization (WHO) released global standards in 2023 to guide TB programs, including a mandate that all people with presumptive TB receive a rapid molecular test as their initial diagnostic exam.(2) Despite being available for over 15 years,(3) WHO-recommended low complexity nucleic acid amplification tests (NAATs) were the initial test for only 54% of the people affected by TB in 2024.(1) This shortfall is partly due to the cost of NAATs and their associated equipment; despite price reductions for available tests, the current price point of around USD 8 per test (1, 4, 5) remains too expensive for most National TB Programs (NTPs) to provide universal coverage. This is exacerbated by the infrastructure needed in health facilities, supply chain disruptions, and reduced diagnostics budgets caused by recent cuts in external global health funding.(6)

Building on investments made to address the COVID-19 pandemic, the development of novel swab-based testing platforms has the potential to cause significant disruption in the TB diagnostic market.(7, 8) New swab-based testing platforms fall under a near point of care (NPOC) NAAT test category, hereafter referred to as NPOC, that has different performance targets based on the sample provided (sputum or non-sputum samples). In March 2026, WHO recommended the use of NPOCs on sputum specimens to increase access to testing, as well as with tongue swabs for adults and adolescents who cannot produce sputum(9).

At the time of writing, three diagnostic accuracy studies documented the performance of the first NPOC test to be included in the WHO recommendations – Pluslife’s MiniDock MTB (MiniDock MTB) (Guangzhou Pluslife Biotech, China). Steadman *et al* reported results from a multi-center study, conducted in outpatient health centers in India, Uganda, and Viet Nam.(8) Mbuli *et al* reported results from a study in Cameroon which recruited participants from both health facilities and the community.(10) Yerlikaya *et al* reported results from a study in outpatient health centers across seven countries.(11) All studies found promising results on swab-based platforms. Results reported sensitivities of MiniDock MTB on swabbed sputum of 91.1% (95% CI: 82.1-95.9%), 86% (95% CI: 79-91%), and 85.7% (95% CI: 83-98%) among culture-positive individuals. This approached the sensitivity of the Xpert MTB/RIF Ultra (Xpert-Ultra) assay (Cepheid, USA) in all studies. MiniDock MTB also showed improved sensitivity compared to smear microscopy for both sputum and tongue swabs.(8, 10, 11)

As well as encouraging performance from initial studies, the MiniDock MTB offers other practical benefits compared to other NAATs. The MiniDock MTB assay is a fraction of the price, requires less infrastructure enabling decentralized deployment, and is simpler to implement as reported by healthcare workers(10). As NTPs are increasingly constrained by diminished external funding, the low costs of the tests and platforms, coupled with the simplicity and more basic infrastructure requirements, may be attractive to implement (10).

However, the real-world performance of many diagnostic tests, such as the NPOCs, may not mirror the results from highly controlled diagnostic accuracy studies; such studies may recruit participants who have different disease characteristics, resulting in differences in detectability of TB (12, 13). This is known as spectrum bias.(12) As well as sensitivity and specificity, all NPOC accuracy studies reported on the concordance – positive percentage agreement (PPA) – of the swab platforms with Xpert-Ultra, disaggregated by semi-quantitative grade (8, 10, 11). This output serves as an assessment of bacterial burden and returns values of High, Medium, Low, Very Low, and Trace (from highest bacterial burden to lowest) (14, 15). Xpert-Ultra expanded the semi-quantitative grading from its predecessor Xpert MTB/RIF (Xpert) by adding the trace category, improving the sensitivity but reducing the specificity of the test(16). Xpert-Ultra uses the detection of multi-copy targets (IS1081 and IS6110) while Xpert MTB/RIF focused on the detection of a single-copy target (rpoB gene)(17). In the available NPOC literature, there is a decrease in assay performance as semi-quantitative results move from High to Trace, with more Xpert-Ultra positive results being missed by the MiniDock MTB assay, both on sputum and tongue swabs for Very Low and Trace results (8, 10, 11). Therefore, it is important to understand the potential trade-offs between Xpert-Ultra and swab-based NPOC assays. In this study, we use results from the existing diagnostic accuracy studies on MiniDock MTB to model its potential performance in a range of populations using a large multicounty dataset that recruited individuals at different levels of health systems. As a point of comparison, we also use an early evaluation of the Xpert-Ultra assay compared to its predecessor the Xpert assay by Dorman *et al* (16). Therefore, we compare the modeled performance of swab-based NPOC tests to Xpert across populations.

## Methods

This was a modeling study to estimate the utility of MiniDock MTB based on the semi-quantitative grade from Xpert Ultra in different populations using data from Phase 1 of the Start4All study conducted in seven countries between November 2023 and February 2025 (https://clinicaltrials.gov/study/NCT05845112). Detailed methods of Start4All have been described elsewhere.(18) Briefly, participants were recruited consecutively and included: people aged ≥15 years in Bangladesh, Cameroon, Kenya, Nigeria, Viet Nam and ≥18 years in Brazil and Malawi with symptoms suggestive of TB (cough, fever, night sweats, weight loss) attending primary health centers and district hospital facilities in all countries; people aged ≥15 years in community settings with TB symptoms and/or abnormal chest X-ray (CXR) (abnormality score of ≥0.3) detected using qXR computer-aided detection (CAD) software (Qure.ai, Mumbai, India) in Bangladesh, Cameroon, Nigeria and Viet Nam.

The ethics committees in each country, as well as the WHO ethics review committee and the Liverpool School of Tropical Medicine institutional review board, approved the study. Participants (and/or caregivers for minors) provided written informed consent in their preferred language.

For the population in our model, we included all individuals ≥15 years from Start4Allwith positive Xpert Ultra results, including trace as positive, and grouped them into two recruitment populations: people identified with presumptive TB via symptom screening and presenting to either hospitals or primary care facilities (facility-based case finding), and people with presumptive TB via CXR and/or symptom screening in community settings (active case finding). Results were disaggregated by semi-quantitative grade on sputum.

To construct the model, we used published data from three diagnostic accuracy studies of swab-based testing, which documented the performance of MiniDock MTB compared to Xpert Ultra (8, 10, 11). We modeled results of MiniDock MTB on sputum swabs compared to Xpert Ultra on sputum as the primary outcome of interest. This was because the Start4All testing was performed on sputum, and because most people with presumptive TB can produce sputum. As a point of comparison, we also used an early diagnostic accuracy study comparing Xpert to Xpert-Ultra, which reported on the results of the two tests disaggregated by semi-quantitative result (16). This was to compare the performance of the predecessor to Xpert Ultra, which did not include the Trace semi-quantitative grade, with the performance of the NPOC test against Xpert-Ultra. Details of the studies used for the model are outlined in Table 1.

**Table 1:** Source studies used in modeling.

| Study | Population | Setting | Data used |
| --- | --- | --- | --- |
| Steadman *et al.* (2025)<br>Diagnostic accuracy of swab-based molecular tests for tuberculosis using novel near point-of-care platforms: A multi-country evaluation (8) | People 12 years and older with presumptive TB, defined as a new or worsening cough lasting $\geq 2$ weeks and/or a risk factor for TB (HIV infection, self-reported close contact with person(s) with TB, or a history of mining work) plus an abnormal WHO-recommended TB screening test (abnormal CXR or, for people living with HIV [PLHIV], C-reactive protein $>5$ mg/L). | Outpatient health centers in India, Uganda and Viet Nam. | PPA of MiniDock MTB sputum swabs positive with Xpert-Ultra positive, disaggregated by Xpert-Ultra grade.<br>A total of 68 Xpert-Ultra positive, including Trace. |
| Mbuli *et al.* (2025)<br>Diagnostic performance of the Pluslife MiniDock MTB and Molbio MTB Ultima assays to detect tuberculosis from tongue and sputum swabs among outpatients and in active case finding in Cameroon (10) | People aged 15 years and older with symptoms (current cough, fever, night sweats, and/or weight loss) and/or clinical risk factors (diabetes, current smoker) for TB | Outpatient departments at health facilities in Cameroon | PPA of MiniDock MTB sputum swabs positive with Xpert-Ultra positive, disaggregated by Xpert Ultra grade.<br>106 Xpert-Ultra positive, including Trace. |
| | People 15 years and older with a CAD score $\geq 0.3$ (qXR Version 4) | Community-based screening camps in rural Cameroon | PPA of MiniDock MTB sputum swabs positive with Xpert Ultra positive, disaggregated by Xpert-Ultra grade.<br>57 Xpert Ultra positive, including Trace. |
| Yerlikaya *et al.* (2026)<br>Pulmonary Tuberculosis Detection with MiniDock MTB Using Swab Samples (11) | People 12 years of age or older, who had either a new or worsening cough lasting at least 2 weeks or at least one tuberculosis risk factor combined with a positive WHO-recommended tuberculosis screening test (abnormal CXR or, for people living with HIV [PLHIV], C-reactive protein $>5$ mg/L). | Outpatient centers in India, Nigeria, the Philippines, South Africa, Uganda, Viet Nam, and Zambia | PPA of MiniDock MTB sputum swabs positive with Xpert-Ultra positive, disaggregated by Xpert-Ultra grade.<br>227 Xpert-Ultra positive, including Trace. |
| Dorman *et al.* (2018)<br>Xpert MTB/RIF Ultra for detection of Mycobacterium tuberculosis and rifampicin resistance: a prospective multicentre diagnostic accuracy study (16) | Adults with TB symptoms | Primary health-care centers and hospitals in eight countries: South Africa; Uganda; Kenya; India; China; Georgia; Belarus; and Brazil. | PPA of Xpert with Xpert-Ultra positive on culture-positive individuals.<br>408 Xpert-Ultra positive, including Trace. |

For the MiniDock MTB performance model, we pooled the reported PPA between MiniDock MTB and Xpert Ultra, disaggregated by semi-quantitative grade reported in the studies by Steadman et al., Mbuli et al., and Yerlikaya et al.(8, 10, 11) The PPAs were pooled using a random effects meta-analysis with restricted maximum likelihood (REML) estimation to derive a single pooled PPA per grade and overall. For the Xpert performance model, we calculated the PPA between Xpert and Xpert Ultra disaggregated by semi-quantitative grade to model performance (Dorman). These PPAs were then applied to the Start4All population to estimate how many positive Xpert-Ultra test results the MiniDock MTB and Xpert may each have missed.

The PPA for each model was applied to the semi-quantitative distribution for positive individual Xpert-Ultra results, both for total recruitment data and disaggregated by facility and community screening to model the performance of the tests in these distinct populations. We used two approaches for the overall predicted positivity and compared results. The first was to use the overall estimate of PPA for each test (MiniDock MTB and Xpert) and apply this to the total number of people with a positve Xpert-Ultra result to predict the overall number detected with each test. The second was to apply the pooled PPA at each semi-quantitative grade to the number of positive Xpert Ultra test results within that grade to account for spectrum bias, applying a version of the spectrum-standardized framework described by Schumacher (13). The predicted number detected within each semi-quantitative grade was then summed to get the total number detected with each test. The two results were then compared. The two approaches are outlined in the equations below, where n is the number of positive Xpert-Ultra test results in the Start4All dataset.

*Approach 1: Single value model*

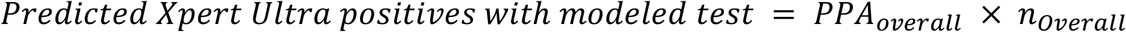

*Approach 2: Semiquantitative grades model*

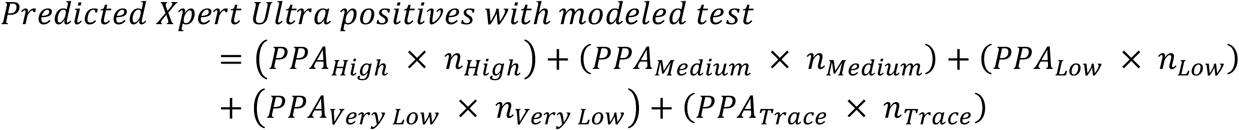

## Results

In the Start4All dataset, there were 1,248 participants with positive Xpert-Ultra results among 13,896 adults across seven countries. Among these, 1,033 (82.8%) were from facility-based recruitment and 215 (17.2%) from active case finiding. Overall, 367 (29.4%) of those had High, 239 (19.2%) had Medium, 317 (25.4%) had Low, 140 (11.2%) had Very Low, and 185 (14.8%) had Trace semi-quantitative grade. The proportions of High, Medium and Low grades were higher in facility-based recruitment compared to active case finding at 30.8% vs 22.8%, 19.7% vs 16.3%, and 26.1% vs 21.9%, respectively. Conversely, the proportions of Very Low and Trace results were higher in active case finding at 14.9% vs 10.5% and 24.2% vs. 12.9% (Table 2). Other participant characteristics are shown in Table 2.

Figure 1 shows the proportion of each semi-quantitative grade across all four studies used for modeling and the Start4All dataset. The highest proportion of Xpert Ultra positives is in the High semi-quantitative category for all MiniDock MTB diagnostic accuracy studies at 35.3%, 37.4%, and 38.3%. However, the proportion of Very Low and Trace results from Mbuli *et al*. is higher than for Steadman *et al*. and Yerlikaya *et al*. While both the multi-site studies only took place in health facilities, the Cameroon study took place in both facility and community settings. For the Dorman *et al*. study, the highest proportion was in the Medium semi-quantitative grade at 57.4%. The lowest proportion was in the Very Low (8.8%) and Trace (3.2%) categories, consistent with the other studies. Full demographic details of each population can be found in the original papers (8, 10, 11).

**Table 2:** Participant characteristics among Xpert Ultra positives from the Start4All study.

|  | Active Case Finding (N=215) | Facility (N=1,033) | Overall (N=1,248) |
| --- | --- | --- | --- |
|  | N (%) | N (%) | N (%) |
| **Age** |  |  |  |
| Median [Q1, Q3] * | 42 [27, 52] | 38 [28, 51] | 39 [28, 52] |
| **Sex** |  |  |  |
| Female | 71 (33.0%) | 335 (32.4%) | 406 (32.5%) |
| Male | 144 (67.0%) | 698 (67.6%) | 842 (67.5%) |
| **HIV status** |  |  |  |
| Negative | 102 (47.4%) | 750 (72.6%) | 852 (68.3%) |
| Positive | 4 (1.9%) | 129 (12.5%) | 133 (10.7%) |
| Unknown | 109 (50.7%) | 154 (14.9%) | 263 (21.1%) |
| **TB symptoms, any from WHO-recommended Four-Symptom Screen (W4SS)** |  |  |  |
| Yes | 191 (88.8%) | 1030 (99.7%) | 1221 (97.8%) |
| No | 24 (11.2%) | 3 (0.3%) | 27 (2.2%) |
| **Xpert-Ultra semi-quantitative grade** |  |  |  |
| High | 49 (22.8%) | 318 (30.8%) | 367 (29.4%) |
| Medium | 35 (16.3%) | 204 (19.7%) | 239 (19.2%) |
| Low | 47 (21.9%) | 270 (26.1%) | 317 (25.4%) |
| Very Low | 32 (14.9%) | 108 (10.5%) | 140 (11.2%) |
| Trace | 52 (24.2%) | 133 (12.9%) | 185 (14.8%) |

\*Q1, Q3: first and third quartiles (25<sup>th</sup> and 75<sup>th</sup> percentiles)

**Figure 1.**
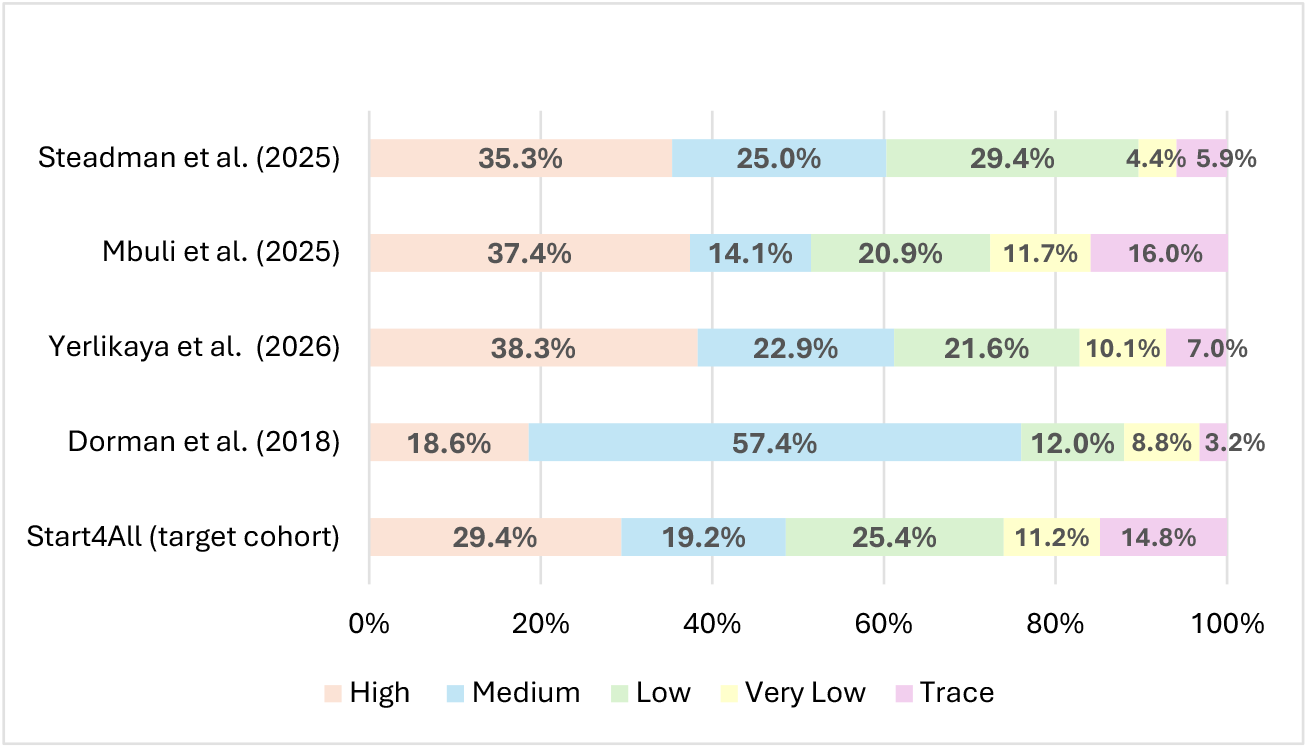
Distribution of semi-quantitative Xpert Ultra grades in studies.

The combined PPA of MiniDock MTB sputum swab and the PPA of Xpert with Xpert-Ultra are shown in Table 3, both overall and disaggregated by semi-quantitative grade. The three MiniDock MTB studies reported a combined overall PPA of 84.8% (67.1-93.8%). The PPA for the individual study populations by Xpert-Ultra grade can be found in the original papers.(8, 10, 11) While the PPA with MiniDock MTB is relatively high for High, Medium, and Low grades, it drops in Very Low and Trace grades at 56.5% (29.8-79.9%) and 33.8% (21.3-48.9%), respectively. Using the results from Dorman *et al*., the overall concordance of Xpert with Xpert Ultra is high at 93.1% (90.3-95.2%); however, only 66.7% (50.3-79.8%) and 23.1% (8.2-50.3%) of Very Low and Trace grades, respectively, were detected as positive by Xpert (16).

**Table 3:** MiniDock MTB sputum swab and Xpert PPA with sputum Xpert-Ultra.

| Xpert-Ultra semi-quantitative grade | Pooled Positive Percentage Agreement from Steadman, Mbuli, and Yerlikaya for MiniDock MTB | Positive Percentage Agreement from Dorman et al. for Xpert |
| --- | --- | --- |
| High | 95.9% (87.8-98.7%) | 100% (95.2-100%) |
| Medium | 97.1% (90.5-99.2%) | 100% (98.4-100%) |
| Low | 88.5% (80.4-93.6%) | 87.8% (75.8-94.3%) |
| Very Low | 56.5% (29.8-79.9%) | 66.7% (50.3-79.8%) |
| Trace | 33.8% (21.3-48.9%) | 23.1% (8.2-50.3%) |
| Overall | 84.8% (67.1%-93.8%) | 93.1% (90.3-95.2%) |

When applying the model in both approaches, results from each cohort show that using the single value overestimates the number of positives which the MiniDock MTB assay would detect in the Start4All dataset; however, the extent of this overestimation varies by population analyzed (Table 4, Figure 2). When applied to the total Start4All dataset, the single model estimate was slightly higher than the individual semi-quantitative model results, with a predicted 1,058 (84.8%) and 1,007 (80.7%) individual positive Xpert-Ultra tests detected, respectively. Facility-based screening results were closer at 876 (84.8%) and 848 (82.1%), respectively. However, the most marked overestimation was in the active case finding cohort, where the single value model (84.8% [95% CI: 67.1-93.8%]) overestimated performance by 10.8% compared to using the individual semi-quantitative grade model (74.0% [61.9-82.8%]).

**Table 4:** Applying MiniDock MTB Predicted Positivity to Start4All Data.

| Xpert-Ultra semi-quantitative grade | Predicted MiniDock MTB Performance | Start4All total recruitment |  |  | Start4All facility data |  |  | Start4All active case finding data |  |  |
| --- | --- | --- | --- | --- | --- | --- | --- | --- | --- | --- |
|  |  | Individual Xpert-Ultra positive | Single value model | Semi-quantitative grade model | Individual Xpert-Ultra positive | Single value model | Semi-quantitative grade model | Individual Xpert-Ultra positive | Single value model | Semi-quantitative grade model |
| **High** | 95.9%<br>(87.8-98.7%) | 367 | - | 352 | 318 | - | 305 | 49 | - | 47 |
| **Medium** | 97.1%<br>(90.5-99.2%) | 239 | - | 232 | 204 | - | 198 | 35 | - | 34 |
| **Low** | 88.5%<br>(80.4-93.6%) | 317 | - | 281 | 270 | - | 239 | 47 | - | 42 |
| **Very Low** | 56.5%<br>(29.8-79.9%) | 140 | - | 79 | 108 | - | 61 | 32 | - | 18 |
| **Trace** | 33.8%<br>(21.3-48.9%) | 185 | - | 63 | 133 | - | 45 | 52 | - | 18 |
| **Overall** | 84.8%<br>(67.1-93.8%) | 1,248 | 1,058 (84.8%<br>[67.1-93.8%]) | 1,007 (80.7%<br>[70.1-88.0%]) | 1,033 | 876 (84.8%<br>[67.1-93.8%]) | 848 (82.1%<br>[71.8-89.1%]) | 215 | 182 (84.8%<br>[67.1-93.8%]) | 159 (74.0%<br>[61.9-82.8%]) |

**Figure 2.**
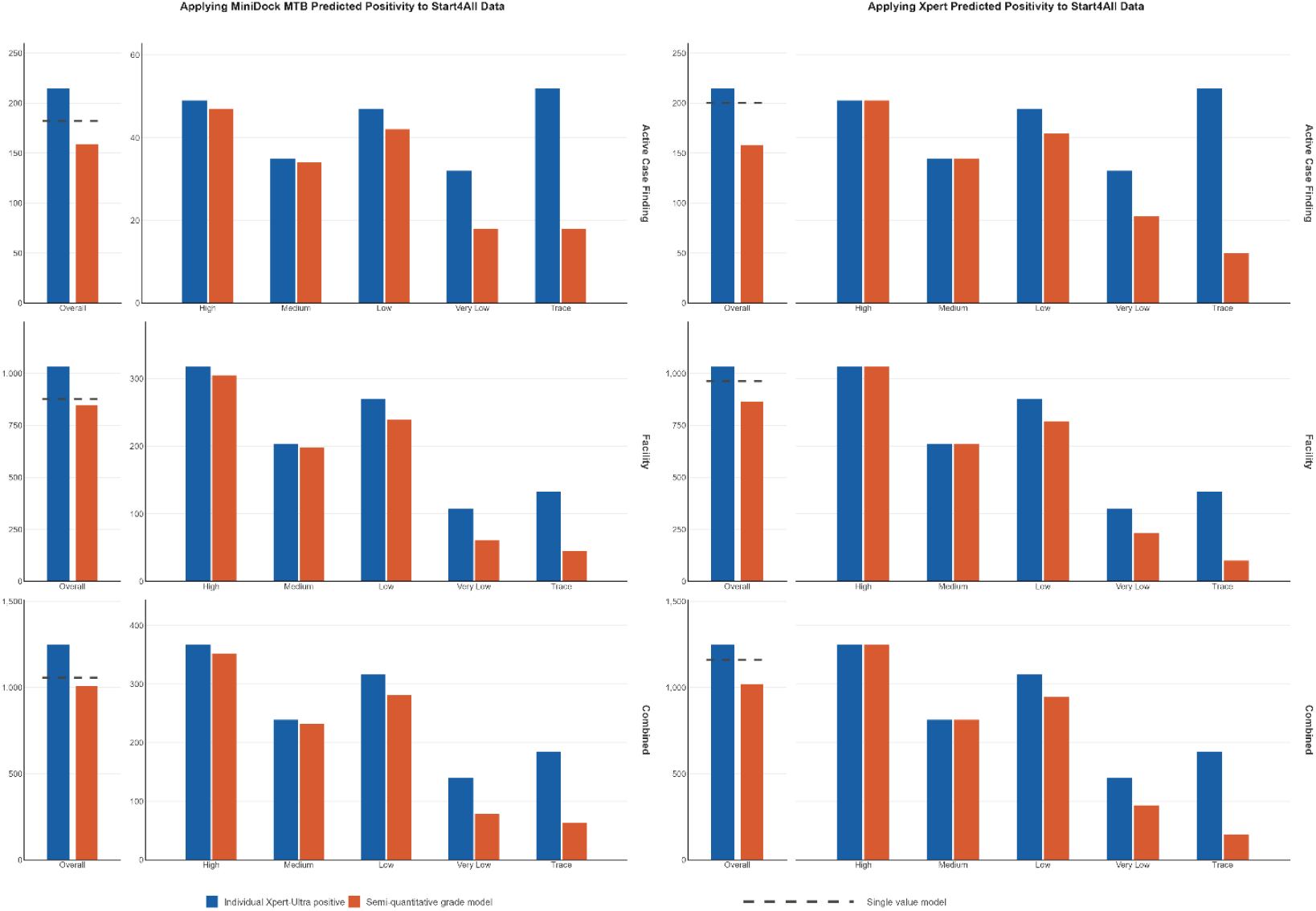
Applying MiniDock MTB and Xpert predicted positivity to Start4All data.

The differences between the single value PPA model and the individual semi-quantitative grade model are even more striking using the Xpert data (Table 5, Figure 2). In the total population, the difference was 11.4% (93.1% vs 81.7%). When disaggregated, facility-based screening had a difference of 9.7% (93.1% v 83.4%), while the difference in active case finding was the largest at 19.1% (93.1 % vs 74.0%). The individual semi-quantitative grades model shows an almost identical performance between Xpert and MiniDock MTB, missing 57 and 56 positive Xpert-Ultra test results, respectively.

**Table 5:** Applying Xpert Predicted Positivity to Start4All Data.

| Xpert-Ultra semi-quantitative grade | Predicted Xpert Performance | Start4All total recruitment |  |  | Start4All facility data |  |  | Start4All active case finding data |  |  |
| --- | --- | --- | --- | --- | --- | --- | --- | --- | --- | --- |
|  |  | Individual Xpert-Ultra positive | Single value model | Semi-quantitative grade model | Individual Xpert-Ultra positive | Single value model | Semi-quantitative grade model | Individual Xpert-Ultra positive | Single value model | Semi-quantitative grade model |
| **High** | 100%<br>(95.2-100%) | 367 | - | 367 | 318 | - | 318 | 49 | - | 49 |
| **Medium** | 100%<br>(98.4-100%) | 239 | - | 239 | 204 | - | 204 | 35 | - | 35 |
| **Low** | 87.8%<br>(75.8-94.3%) | 317 | - | 278 | 270 | - | 237 | 47 | - | 41 |
| **Very Low** | 66.7%<br>(50.3-79.8%) | 140 | - | 93 | 108 | - | 72 | 32 | - | 21 |
| **Trace** | 23.1%<br>(8.2-50.3%) | 185 | - | 43 | 133 | - | 31 | 52 | - | 12 |
| **Overall** | 93.1%<br>(90.3-95.2%) | 1,248 | 1,162 (93.1%<br>[90.3-95.2%]) | 1,020 (81.7%<br>[70.6-88.9%]) | 1,033 | 962 (93.1%<br>[90.3-95.2%]) | 862 (83.4%<br>[74.9-90.0%]) | 215 | 200 (93.0%<br>[90.3-95.2%]) | 158 (73.5%<br>[63.7-83.7%]) |

## Discussion

Our results demonstrate the spectrum bias effect, where single point estimates from diagnostic accuracy studies can overestimate real-world test performance, due to differences in population and distribution of burden of disease. This applied to both MiniDock MTB and Xpert and highlights the importance of evaluating performance across the range of disease burden characteristics, represented here by semi-quantitative grades, rather than a single value. Our study suggests that MiniDock MTB sputum swabs might perform comparably to Xpert in detecting people with TB, when performance is assessed using semi-quantitative grades rather than a single pooled PPA value. While using a single study estimate would suggest that Xpert performs better and would miss fewer people confirmed with TB with Xpert-Ultra, taking the semi-quantitative grades into account shows minimal differences. Using a singular PPA value also hides the varying performance of the assays in different screening contexts; when using the semi-quantitative grade values in the model, both tests are expected to miss more people who are TB positive on Xpert-Ultra in active case finding rather than facility settings.

Our results corroborate Kendall *et al*.’s argument that simple numerical estimates of accuracy, such as sensitivity and specificity, do not always show the full picture of who with TB is detected and missed (12, 13) Characteristics of TB, such as bacterial load, lung parenchymal changes (such as cavitation), or localization versus dissemination of the bacterium, differ from person to person, and this influences whether they are detected by any given assay (12, 13, 19). The majority of diagnostic accuracy studies tend to take place in facilities, often in reference laboratories, among care-seeking or symptomatic people who do not reflect general populations, such as those at community screening events or lower-level health facilities and can differ from programmatic profiles due to strict enrolment criteria. In order to aid the translation of these results to different populations, characterizing TB disease across dimensions, such as Xpert-Ultra semi-quantitative grades, can help to predict how assays may perform in alternative settings (12, 13). Our results demonstrate how this breakdown across a spectrum of disease can dramatically change the number of people with TB predicted to be detected and missed when translating results from diagnostic accuracy studies.

There was a higher proportion of High grade positive Xpert-Ultra test results in facilities compared to active case finding from both Start4All and the study in Cameroon, which suggests a population with a higher bacteriological load.(20-22) Similarly, studies across several settings and countries have shown that more people with TB found through community-based efforts have smear-negative TB compared to people detected through facility-based screening.(23-25)

Our findings raise questions about where and for whom novel NPOC tests may provide the most benefit. As outlined above, in terms of yield, it may be best to use MiniDock MTB in facilities rather than in the community, where higher concordance with Xpert-Ultra would be expected. Similarly, using MiniDock MTB at lower levels of the care system or in community settings carries a risk of missing substantial numbers of people with TB, as it is likely that a higher proportion of these populations will have a lower bacterial burden.(26, 27) Bacillary loads also tend to be lower in certain vulnerable groups, such as PLHIV and children (16, 28, 29).

On the other hand, our study suggests that MiniDock MTB performance may be similar to Xpert in the lower grades, with a higher proportion of people with TB missed in the Very low and Trace grades. While Xpert has now been phased out and replaced by Xpert-Ultra, MiniDock MTB sputum swabs offer a similar-performing alternative at a fraction of the price. MiniDock MTB is currently less than US$4 per test, compared to US$8 for Xpert Ultra and US7.90 for Truenat, with a testing platform more than twenty times less expensive than a four-module GeneXpert system and TrueLab Quatro (4, 5, 8, 10, 30).

MiniDock MTB also has the advantage of being a simpler test to perform than Xpert Ultra, requiring fewer trained staff or lab capacity, with healthcare workers in Cameroon reporting that running the tests was an easy procedure with limited training required.(10) The platform can also be used directly in the field, in contrast to Xpert Ultra, with a solar power supply and results available in less than 30 minutes, compared to around 80 minutes for Xpert-Ultra (10, 17). Therefore, employing this technology at the community level could still help to detect people with TB who otherwise would not have access to testing, due to a lack of lab capacity or equipment. However, it is important to remember limitations with MiniDock MTB for throughput, as only one test can be run at a time in the standard platform, compared to multiple tests for Xpert-Ultra.

There are discussions around whether people with Trace results should be initiated on treatment, as it may indicate previous TB. Some studies suggest that monitoring and retesting Trace results can frequently result in a higher-grade confirmed positive test (28). For example, in a study during a community-based active case finding campaign in Ho Chi Minh city, Viet Nam, almost 40% of all positive Xpert-Ultra test results were within the Trace grade. Of the 85% of these retested, 46% were positive, and represented nearly 20% of all people with TB detected.(31) Similarly, a study in Uganda and South Africa found that almost half of all trace positive individuals not initially started on treatment were diagnosed with TB within three months.(32) As swabs return a binary result, the ability to monitor individuals with lower bacillary loads is not possible, and thus, people with TB may be missed.

Additional tests, such as CXR with CAD software to monitor individuals who exhibit abnormal CXRs but have a negative test, may offer potential opportunities,(12) although the practical implementation of monitoring and retesting individuals with Trace results presents challenges, particularly in community-based settings where re-engagement with care can be difficult.

This study has several limitations. MiniDock MTB has not been widely used yet, and as such, the data we have used as the basis of our MiniDock MTB model – from Steadman *et al*, Mbuli *et al*, and Yerlikaya *et al* – are from relatively small population sizes and thus may not fully reflect the performance of MiniDock MTB across diverse settings. We also only included data on the use of these swabs on sputum specimens, which misses tongue-swab specimens, a key use-case for NPOC tests. The Start4All study is also not representative of programmatic settings; however, our study aims to demonstrate the potential differences in results when different populations are tested. There were also meaningful differences between the populations used to derive the model PPAs and the Start4All population, and MiniDock MTB was not tested directly alongside Xpert Ultra or Xpert on the same samples, meaning the comparison is modeled rather than head-to-head. As such, MiniDock MTB may have performed differently had it been implemented within Start4All. For the Xpert model, we only used data from one study (Dorman *et al*.). While there are other additional diagnostic accuracy studies, such as in a Cochrane review, none reported PPA per grade and could therefore not be integrated into the model (33).

## Conclusion

This study demonstrates the differential modeled performance of MiniDock MTB sputum swabs across screening contexts and how it compares to Xpert. The study also serves as a demonstration of the need to consider evaluating new diagnostics by taking into account the spectrum of TB disease and how this influences the translation of tests across different settings (12). Further prospective studies, including direct comparisons of MiniDock MTB and other swab-based technologies across different specimens and populations, are needed to confirm these findings and inform programmatic rollout.

## Data Availability

All individual participant data collected as part of the Start4All study, after deidentification, will be made available upon reasonable request. The materials to be shared include the individual participant dataset together with the Study Protocol, Statistical Analysis Plan, Informed Consent Form, Clinical Study Report, and analytic code. Data will be available immediately following publication, with no end date. Access will be granted to anyone who wishes to use the data for any purpose. The data and accompanying documents will be available through a controlled access process.

